# Smartphone videos of the sit-to-stand test predict osteoarthritis and health outcomes in a nationwide study

**DOI:** 10.1101/2022.09.29.22280368

**Authors:** Melissa Boswell, Łukasz Kidziński, Jennifer Hicks, Scott Uhlrich, Antoine Falisse, Scott Delp

**Author notes:** Authors contributed equally to this work.

## Abstract

Physical function decline due to aging or disease can be assessed with quantitative motion analysis, but this currently requires expensive laboratory equipment. We introduce a self-guided quantitative motion analysis of the widely used five-repetition sit-to-stand test using a smartphone. Across 35 US states, 405 participants recorded a video performing the test in their homes. We found novel relationships not detectable in a clinical implementation of this test. Trunk angle during the sit-to-stand transition was greater in individuals with osteoarthritis and differed across ethnicities. In individuals 50 years of age or older, those with greater trunk angular acceleration had a higher mental health score. We also detected known associations between longer time to complete the five repetitions and lower physical health scores, higher BMI, and older age. Our findings demonstrate that at-home movement analysis goes beyond established clinical metrics to provide objective and inexpensive digital outcome metrics for nationwide studies.

## Main Text

Physical function profoundly impacts an individual’s quality of life^1^, as evidenced by the diminishing functional health observed with aging^2^ and diseases such as osteoarthritis^3^. Declining physical function in older adults is associated with increased falls, medical diagnoses, doctor visits, medications, and days spent in a hospital^2^. The time required to complete five repetitions of the sit-to-stand (STS) transition, as measured by a stopwatch, is widely used to evaluate physical function. In-lab studies indicate that automated timing is more sensitive in detecting physical health status than manual measurement^4,5^, and kinematic measures are more sensitive than timing alone^6–8^. However, quantifying human movement traditionally requires an expensive motion-capture system and experienced laboratory personnel, severely restricting scalability and access.

The rapid increase in smartphone availability^9^ and recent developments in video-based human pose estimation algorithms^10–13^ may allow automated motion analysis using two-dimensional (2D) video recorded with a smartphone^14,15^. Yet, to date, studies analyzing motion from smartphone videos have been carried out in a clinical^14^ or laboratory setting^15^. In a recent home-based study, STS test time extracted from skeletal motion data from the Microsoft Kinect color camera and depth sensor correlated with participants’ laboratory-based time^16^. This study supports the feasibility of unsupervised at-home tests; however, research staff trained participants to conduct the test in their homes, and the requirement of owning a Kinect inhibits broad adoption. It remains unclear whether pose estimation from self-recorded smartphone video can quantify movement with sufficient accuracy to predict health and physical function.

Here, we examine whether at-home smartphone videos of the STS test predict clinically relevant health measures. To do this, we developed an online tool to capture and automatically analyze self-collected at-home videos of the five-repetition STS test (Fig.1A and Extended Data Fig. 1). This tool also collected demographic and health data via surveys. We deployed the tool in a nationwide study and examined if the data reproduced relationships from previous laboratory studies. To assess the accuracy of our home-based system, we compared the STS parameters extracted from our web application with those calculated from a laboratory motion capture camera system. We then examined whether quantitative STS parameters related to measures of demographics, physical health, mental health, and knee or hip osteoarthritis diagnosis. Osteoarthritis was the primary health condition we evaluated due to its widespread prevalence^17^ and well-documented effect on lower body strength^18^ and altered STS kinematics^6,19^.

**Fig. 1.**
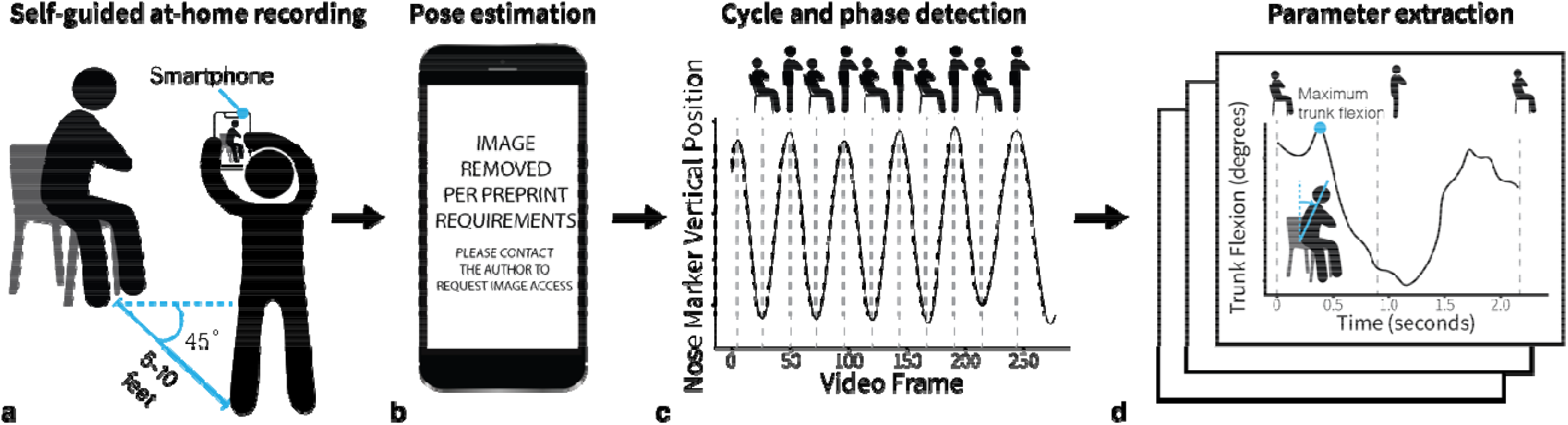
An overview of our web application to collect and analyze movement data. **A)** Participants perform the five-repetition sit-to-stand test while an untrained individual records the test using only a smartphone or tablet from a 45-degree angle to capture a combined sagittal and frontal view. **B)** The video is uploaded to the cloud and a computer vision algorithm, OpenPose^12^, computes body keypoints throughout the movement. **C)** Our tool computes the key transitions in each STS cycle (i.e., as the participant rises from the chair and returns to sitting). **D)** Our algorithms compute the total time to complete the test and several important biomechanical parameters, like trunk angle (see Methods for details).

From 493 total videos submitted, 405 videos across 35 US states were used in the final analysis (Extended Data Fig. 2). Participant characteristics are described in Extended Data Table 1. Our study had nearly 35 times the number of participants of traditional biomechanical studies (where the median sample size is 14.5^20^) with minimal researcher time and resources required.

We first examined if our tool could reproduce the results of laboratory and clinic-based assessments. We found that a larger maximum trunk angle was associated with a diagnosis of osteoarthritis (R=0.18, p<0.001; Extended Data Table 2), even when controlling for age, sex, BMI, and STS time (β=0.029, 95% CI=[0.006, 0.052], p=0.015; Extended Data Table 3). The difference in trunk angle between groups in our study (5.8 degrees) was smaller than the difference in trunk flexion reported by Turcot et al. (9.0 degrees), which is expected since the prior study only included people with advanced knee osteoarthritis. Additionally, Turcot et al. measured trunk flexion purely in the sagittal plane^6^, while the trunk angle obtained from a 45-degree angle in our study is affected by movement in both the frontal and sagittal planes. Previous lab-based studies have also found that individuals with knee osteoarthritis adopt a larger trunk flexion angle and greater lateral trunk lean on the contralateral side during the STS transition to reduce the knee joint moment, joint contact forces, or pain^6^, or to compensate for weak knee extensor muscles^18,21^. Our smartphone-based tool was able to capture this kinematic compensation (Fig. 2A). STS time was associated with osteoarthritis (R=0.18, p=0.001; Extended Data Table 2), but was no longer a significant predictor of osteoarthritis status when controlling for age, sex, and BMI (p=0.847). Thus, while time to complete the task is related to other measures, kinematics appear to be a more specific and sensitive measure of health and functioning. We found moderate to strong associations between STS parameters extracted from our web application and from the same or the closest analogs from motion capture (R=0.997, R=0.583, R=0.702, and R=0.556 for STS time, maximum trunk angle from video vs. lumbar flexion from motion capture, maximum trunk angle from video vs. lumbar bending from motion capture, and maximum trunk angular acceleration from video vs. lumbar flexion acceleration from motion capture, respectively; see Methods).

**Fig. 2.**
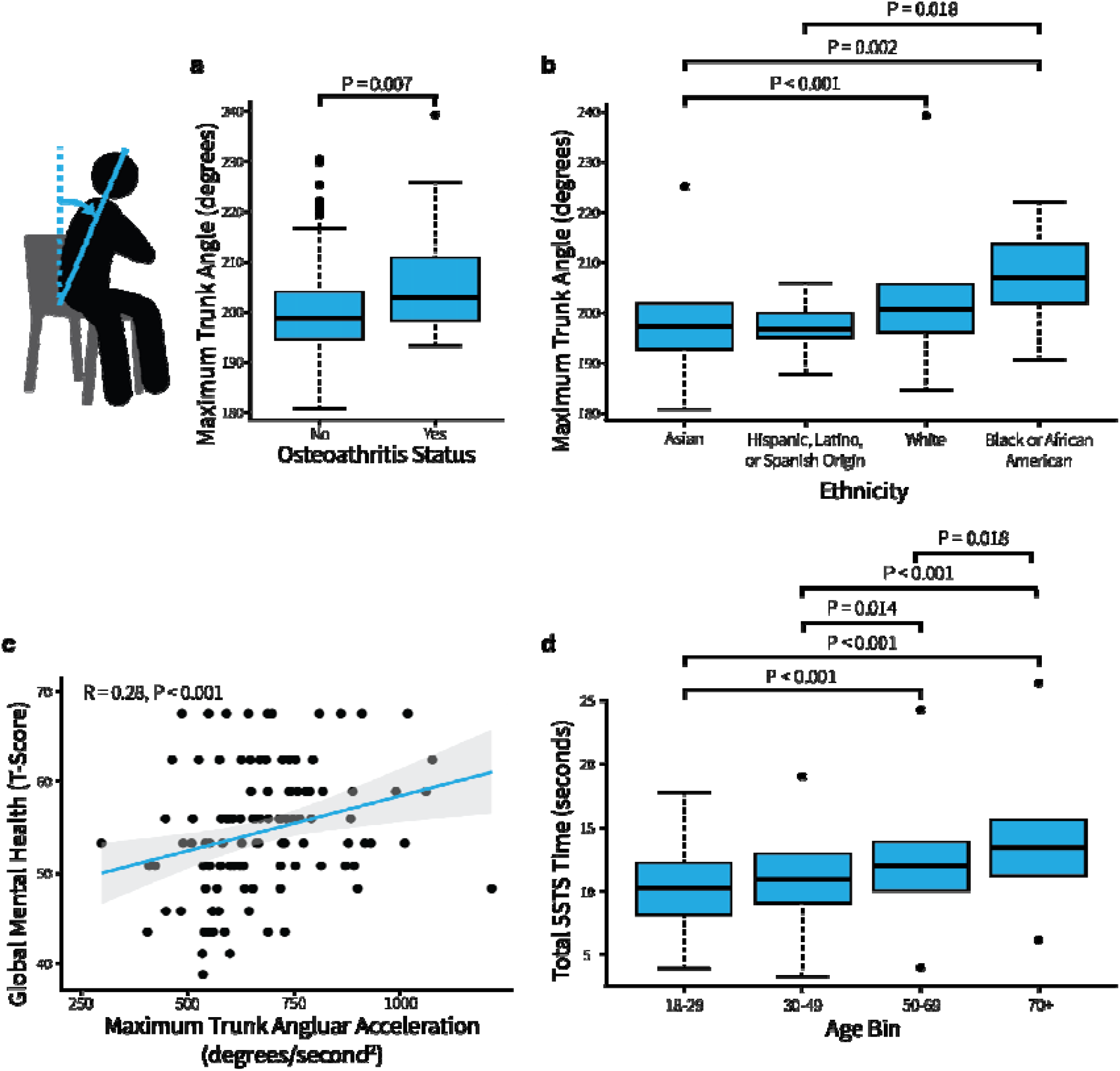
Relationships between sit-to-stand parameters and survey measures. **A)** Trunk angle is larger in patients with hip or knee osteoarthritis, determined from a Pearson correlation test adjusted to control for the false discovery rate. **B)** Trunk angle differs across race and ethnicity, determined from a Dunn’s test with multiple comparison p-values adjusted to control for the false discovery rate. **C)** Greater trunk angular acceleration is associated with a higher mental health score, determined from a Pearson correlation adjusted to control for the false discovery rate with all 21 comparisons. **D)** Test completion times increase with older age, as determined by a t-test. In the box-and-whisker plots, the top and bottom lines of the boxes (hinges) are the first and third quartiles, respectively. The horizontal line is the median, and the whiskers extend from each hinge to the largest value no further than 1.5 times the interquartile range to the respective hinge. In the scatter plot, the grey shading around the blue regression line represents the confidence interval in the scatter plot.

Our smartphone-based tool also reproduced the significant positive associations between STS time and health, age, and BMI found in prior lab-based studies^4,23,24^. In particular, a longer time to complete the STS test was associated with a lower physical health score (R=-0.20, p<0.001), a higher BMI (R=0.20, p<0.001), and older age (R=0.35, p<0.001; Fig. 2C and Extended Data Table 2). Further, time was a predictor of physical health (β=-0.938, 95% CI=[-1.610, -0.237], p=0.006) when controlling for age, sex, and BMI. All other relationships evaluated were not significant (Extended Data Table 2). Compared to reference STS test times, the average test time in our study was longer (11.4±3.4 seconds vs. 7.5±2.4 seconds reported by Bohannon et al.^24^). A similar minimum (4.3 vs. 3.9^24^ seconds) but larger maximum (32.9 vs. 17.6^24^ seconds) indicates greater variation in performance, possibly due to the lack of feedback and test training^25^.

We next explored relationships between STS parameters across varying ethnic and racial groups and mental health. Maximum trunk angle differed across racial and ethnic groups (p<0.001; Fig. 2B; Extended Data Table 4). In a comparison between the two largest ethnic groups, white (N=243) vs. Asian (N=103), differences in trunk angle remained significant when controlling for age, sex, BMI, and physical health (β=-0.084, 95% CI=[-0.130, -0.038], p<0.001; Extended Data Table 5). While racial and ethnic disparities exist in the incidence and outcomes of musculoskeletal disease^26^, race and ethnicity are rarely examined in biomechanical studies due to the typically small study samples. Similar to the conclusions of Hill et al. who found racial differences in gait mechanics^27^, our findings suggest that we should not assume biomechanical similarity between different racial and ethnic groups.

Since STS tests are most commonly performed in older adults, we also performed an exploratory subgroup analysis between STS parameters and physical and mental health in the 106 individuals 50 years of age or older. We found that greater maximum forward trunk angular acceleration was associated with a higher mental health score (R=0.28, p=0.012; Fig. 2D; Extended Data Table 6), which remained significant when controlling for age, sex, BMI, and time (β=1.705, 95% CI=[0.376, 3.034], p=0.012; Extended Data Table 7). Psychological studies typically require larger sample sizes than biomechanical studies to determine significant results; therefore, few studies have evaluated the relationships between biomechanics and mental health. The large-scale of these at-home tests could allow further exploration of these relationships and, potentially, enable the use of one’s motion as an objective measure of mental health status.

Participants considered the protocol very easy (see Methods), suggesting real-world adherence to our app would be high^28^, but limitations remain. One key limitation is the inconsistency in STS test performance and environment across participants. For example, participants used varying types and heights of chairs and foot and arm positions, which can influence the STS movement^25^. Our study’s 2D joint angle projections were likely affected by camera recording angle and height differences, but our large number of participants helped overcome this variability. Improved user interfaces could more consistently guide a participant to the correct position relative to the camera and reduce this variability. Future advances in 3D pose estimation could mitigate camera position issues and be integrated with a musculoskeletal model to obtain kinetic measures such as joint loading^22^. Another limitation was the error of the pose-estimation algorithm when predicting joint locations (particularly the hip) for individuals with loose-fitting clothing, such as skirts or sweatpants, or higher BMIs. Where apparent, these videos were removed, but the errors with hip location estimation may have still influenced our results, particularly trunk kinematics. Pose-estimation algorithms are often tested on large datasets of individuals performing a range of activities^29,30^; digital tools meant for health evaluations may benefit from additional model training with a diverse sample of participants (i.e., varying BMIs), particularly those with movement conditions like osteoarthritis, performing the activity of interest.

In summary, we developed a digital tool to automatically measure STS times and kinematics from at-home videos, deployed it in a nationwide study, and found that measurements from at-home videos are sensitive enough to predict physical health and osteoarthritis. The consistency of this study’s results with lab-based studies, including the relationship between trunk angle and osteoarthritis presence, and its accessibility as an open-source online tool support its use by researchers and clinicians to leverage biomechanics for at-home monitoring of physical functioning at an unprecedented scale. Further, with a large pool of participants, we discovered relationships between biomechanics and ethnicity and race, as well as biomechanics and mental health. Our web app, data set, source code, and processing code are freely available online, enabling other researchers to use and adapt our tools and explore our dataset for new research questions. For example, researchers could adapt our web app to analyze other variations of the STS test or different functional tests so that in the future, it may be possible to conduct an entire battery of functional tests at home. Our tool can also analyze previously collected video data, opening the door to answering a multitude of new research questions without any additional data collection. Our study demonstrates the feasibility of mobile, cheap, and easy-to-use tools for biomechanical assessment of functional tests that may allow anyone with a smartphone to objectively measure their health and physical functioning.

## Methods

### Participants and procedures

#### Participants

Across 35 US states, 493 participants (age: mean = 37.5 years, range = (18, 96); sex: 54% female) successfully completed the entirely self-guided study. Individuals were qualified to participate if they currently resided in the United States, were of at least 18 years of age, had gotten up and down from a chair in the past week, felt safe standing up from a chair without the use of their arms, indicated that another person was present to monitor and record their test, and answered “No” to all questions of risk in the Physical Activity Readiness Questionnaire for Everyone (2020 PAR-Q+)^31^. In the sample of individuals used in the final analysis (N=405; participant exclusion described in the “Data Cleaning” section), the mean age was 37.3 ± 17.8 years, ranging from 18 to 96 years, and 53% were female (Extended Data Table 1). To test for differences in age, gender, and BMI between participants included vs. excluded from the analysis, we calculated the standardized mean difference (SMD)^32^. We chose an SMD of less than 0.1 to indicate a negligible difference, a threshold recommended to determine imbalance^33^. There were no differences in age or BMI between participants included and participants excluded from the final analysis (SMD (95% Confidence Interval (CI)) = 0.07 (−0.16; 0.30) and SMD (95% CI) = 0.05 (−0.17; 0.28), respectively); however, there was a larger proportion of female participants in the entire sample than those included in the final analysis (58% versus 53%, respectively; SMD (95% CI) = 0.10 (−0.13; 0.33)).

Our team recruited participants via social media posts, fliers, word of mouth, and other study participant pools. By leveraging research studies focusing on aging and osteoarthritis, we recruited individuals of older age and with hip and/or knee osteoarthritis. Participants were compensated with a $30 gift card and received a link to their STS test with an overlaid visualization of their motion analysis. We obtained approval for the study from the Stanford University Institutional Review Board (IRB-59455) and digital informed consent from all participants.

#### Procedures

Participants joined our study directly from our website (sit2stand.ai; <u>Extended Data Document 1</u>). After selecting “Join Study,” they were directed to a series of qualification and safety questions. If they qualified, they were presented with a digital consent form. Immediately after providing informed consent, participants were shown a video and written instructions for the STS test. The webpage gave the option to open the individual’s camera to record the test or upload a previously recorded video. After upload, the participant reviewed their video and approved it for submission before being directed to the survey (<u>Extended Data Document 2</u>).

#### Five-repetition STS test

We chose the STS test as it is a frequently used clinical test of physical function. The STS transition is related to the strength and power of the lower limbs^24^, such as knee extension strength^34^. It is one of the most mechanically demanding functional daily activities^35^. Because of this, clinicians and researchers widely use STS transitions to evaluate physical function. Additionally, a recent study tested the feasibility of administering the STS test at home and found that a self-administered, video-guided STS test was suitable for participants of varying ages, body sizes, and activity levels^36^.

In the most common variation of STS transition tests, the five-repetition STS test^37^, an individual moves from sitting in a chair to standing five times in a row as quickly as possible with their arms folded across their chest (Extended Data Fig. 3). Researchers have related the time to complete the STS to age, height^24^, weight^24^, knee extension strength^24^, physical activity level^4^, vitality^16^, anxiety^16^, and pain^16^. STS is also a valid and reliable clinical assessment for various conditions, including arthritis^38^, pulmonary disease^39^, Parkinson’s disease^40^, and degenerative spinal pathologies^41^. Beyond timing, in-lab studies have found that one’s kinematics during an STS task are related to frailty^7^, fall risk^16^, and osteoarthritis status^19^.

### Survey measures

#### Participant characteristics

Participants reported, via survey, their age, sex, gender, height, body weight, ethnicity, education, employment, income, marital status, and state of residence. BMI was calculated from their reported height and weight.

#### Physical and mental health

Overall physical and mental health status was assessed using the PROMIS v.1.2 Global Health Short Form^42^. The Global Health Short Form is a ten-item survey measuring overall physical function, fatigue, pain, emotional distress, and social health in healthy and clinical adult populations. Separate scores were calculated for global physical health (GPH) and global mental health (GMH)^43^.

#### Osteoarthritis status

Participants were asked (yes/no) whether they have a clinical diagnosis of hip or knee osteoarthritis.

### Video analysis

#### Automated pose estimation

We instructed each participant to record a video of the STS test using a smartphone placed or held vertically. We processed all videos using OpenPose (Cao, 2018), a widely used^44^, and high-performing^45,46^ neural network-based software for pose estimation. For each person present in an RGB image, OpenPose returns the 2D position of 25 body landmarks: the nose, neck, and midpoint of the hips, and bilateral shoulders, elbows, wrists, hips, knees, ankles, eyes, ears, first metatarsals, fifth metatarsals, and heels.

From each video, we extracted frames using FFmpeg Version 4.2.4 and ran OpenPose on each video frame. In frames where the algorithm detected multiple people, we only considered the person closest to the camera, defined as the detection with the greatest distance between the feet and nose. Pose estimation processing failed for four videos, which were not included in the final analysis.

#### Pre-processing

We derived the number of frames per second (framerate) for each video using ffprobe software. OpenPose failed to detect the participant’s pose in a small fraction of frames (< 1%). As only a single frame was ever missing in a series, we used linear interpolation to estimate missing keypoint positions in a given frame. We observed high-frequency, low-magnitude noise in the OpenPose output, possibly due to the low resolution of the input for the OpenPose neural network. We found that a 6 Hz, fifth-order, zero-lag, low-pass Butterworth filter (scipy package) was the most robust when comparing low-pass filtering, spline smoothing, and Gaussian smoothing. While we instructed the recorder to record to the right of the participant, for consistency, we horizontally mirrored keypoints in cases where the participants’ left side was closest to the camera. To normalize the data across participants, we divided all coordinates by subject height in pixels, approximated as the 95th percentile of the distance between the right ankle and nose keypoints. For 32 participants, our algorithm detected 3 (N=1), 4 (N=29), or 6 (N=2) STS cycles. We found the average time per cycle for these participants and multiplied it by five.

#### STS parameter extraction

We used the nose marker’s local peaks in the vertical axis to determine the standing and sitting phases. We defined the STS phase as the time between a local minimum and the following local maximum and the stand-to-sit phase as the time between a local maximum and the following local minimum. We calculated the total test time as the time between the first and the last standing positions.

We computed 2D joint angles in the camera projection plane for the right and left sides, including the knee (from the ankle-knee-hip keypoints), hip (from knee-hip-neck keypoints), and ankle (from the first metatarsal-ankle-knee). We defined trunk angle as the angle between a vector from the hip pointing vertically along the camera frame and a vector from the right hip to the neck. To compute marker speeds and joint angular velocities and accelerations, we used discrete derivatives and divided them by the frame rate. We averaged total test metrics across the five stand-to-sit-to-stand cycles and isolated for the sit-to-stand and stand-to-sit phases. Prior to analysis, we hand-selected a limited set of kinematic parameters (i.e., trunk angle and trunk angular acceleration during the sit-to-stand transition) based on previous literature and assessed their associations with the survey measures.

#### Data cleaning

Out of 493 videos submitted, 489 were successfully processed with OpenPose. From this subset, we excluded 84 participants due to the following video recording errors (not mutually exclusive): use of a heavily cushioned chair (n=2); long pause between repetitions (n=29); too close, out of frame, or bodily obstruction (n=25); camera angle was planar (rather than at a 45-degree angle; n=34); use of arms to stand (n=20); large pose-estimation error due to participant wearing a skirt (n=1).

### Statistical analyses

#### Descriptive statistics

Standard descriptive statistics were calculated for participant characteristics, outcome measures, and STS times and kinematics.

#### Associations

We used Pearson correlations to evaluate associations between STS parameters (time, maximum trunk angle during STS, and maximum trunk angular acceleration during STS) and characteristics (age, sex, BMI, and ethnicity) and health measures (physical health, mental health, and osteoarthritis diagnosis). We accounted for multiple comparisons of the Pearson correlations by controlling for the false discovery rate (Benjamini and Hochberg method^47^) with all 21 comparisons. For analyzing associations between STS parameters and physical and mental health in the subsample of participants over the age of 50 (N = 106), we accounted for multiple comparisons with six comparisons.

To compare trunk angles among the four largest ethnic and racial groups, we performed a Kruskal-Wallis test, which accounted for the non-parametric distribution of the smallest two groups. We followed this test with a Post-hoc Dunn’s test with multiple comparison p-values adjusted to control for the false discovery rate. Additionally, we performed a logistic regression with the two largest groups (white vs. Asian), controlling for age, sex, and BMI. Significant associations between kinematic parameters and health measures were further evaluated with linear or logistic regression (for continuous or binary dependent variables, respectively), controlling for age, sex, BMI, and STS time.

### Lab-based motion capture validation

We compared our video-based STS parameters to laboratory measurements from marker-based motion capture.

#### Participants

We collected data from eleven healthy adults (N = 11, 7 female and 4 male; age = 27.7 ± 3.4 [23-35] years; body mass = 67.8 ± 11.4 [54.0-92.9] kg; height = 1.74 ± 0.11 [1.60-1.96] m; mean ± standard deviation [range]). All participants provided written informed consent before participation. The study protocol was approved and overseen by the Institutional Review Board of Stanford University (IRB00000351).

#### Protocol

We measured ground truth kinematics with an eight-camera motion capture system (Motion Analysis Corp., Santa Rosa, CA, USA) that tracked the positions (100Hz) of 31 retroreflective markers placed bilaterally on the 2nd and 5th metatarsal heads, calcanei, medial and lateral malleoli, medial and lateral femoral epicondyles, anterior and posterior superior iliac spines, sternoclavicular joints, acromion processes, medial and lateral epicondyles of the humerus, radial and ulnar styloid processes, and the C7 vertebrae. Twenty additional markers aided in limb tracking. Marker data were filtered using Savitzky-Golay filter with a window size of 0.5s and a 3rd-degree polynomial.

We used OpenSim 4.3^48,49^ to estimate joint kinematics from marker trajectories. We first scaled a musculoskeletal model^50^ to each participant’s anthropometry based on anatomical marker locations from a standing calibration trial using OpenSim’s Scale tool. Then we computed joint kinematics using OpenSim’s Inverse Kinematics tool.

We computed the time to complete the STS test using motion capture and OpenPose. Since the nose marker was not collected in motion capture trials, for comparability, we used the peaks of the pelvis marker in both settings. We compared the video-based test time and kinematic measures (total trunk angle and trunk acceleration) to motion capture test time and the most similar kinematic parameters (lumbar flexion and bending and lumbar flexion acceleration). For these comparisons, we used r statistic, the square root of the coefficient of determination R^2^.

### Participant feedback

#### Participant feedback

Participants rated the difficulty of their participation with the question, “How easy or difficult was it for you to complete the STS test portion of this study (including reading the instructions, performing the test, and uploading the video)?”. An open-ended follow-up question allowed participants to further describe any challenges or general feedback.

On average, participants found completing the study “very easy” to do (4.58 ± 0.77, N = 493; 1 = very difficult and 5 = very easy). A thematic analysis of participant feedback uncovered eight themes related to the experience of participating in the study: The study was enjoyable (e.g., “Really enjoyed being able to participate in something from home, pretty cool!”); the study was easy to do (e.g., “The instructions were clear and [the] platform was easy to use”); participants were curious about the purpose of the study and interpretation of the results (e.g., “Would have been great if you could explain how my responses would help in the study”); the study took longer than expected, particularly the survey portion (e.g., “Survey too long”); participants were confused or had suggestions about the STS instructions (e.g., “The record button followed by more instructions was confusing”); participants were confused or had suggestions about the survey; (e.g., “Wording of questions a little confusing “); participants had technical challenges (e.g., “Instruction video didn’t play”); and participants indicated personal preferences or challenges (e.g., “I prefer a computer to a phone”).

## Data Availability

All data produced are available online at https://github.com/stanfordnmbl/sit2stand-analysis.

https://github.com/stanfordnmbl/sit2stand-analysis

https://sit2stand.ai/

## Acknowledgments

We would like to thank the Stanford Women’s Health Initiative Strong and Healthy for their support in participant recruitment. This work was funded by National Science Foundation Graduate Research Fellowships (grant no. DGE-1656518); the Stanford Catalyst for Collaborative Solutions; the Mobilize Center, which is supported by the National Institute of Biomedical Imaging and Bioengineering (NIBIB) and the Eunice Kennedy Shriver National Institute Of Child Health & Human Development (NICHD) of the National Institutes of Health (NIH) under Grant P41EB027060; and the Center for Reliable Sensor Technology-Based Outcomes for Rehabilitation (RESTORE), which is supported by the Eunice Kennedy Shriver National Institute Of Child Health & Human Development (NICHD) and the National Institute Of Neurological Disorders And Stroke (NINDS) of the National Institutes of Health (NIH) under Grant No. P2CHD101913.

## Data Availability

All data generated in this study are available on GitHub: github.com/stanfordnmbl/motionlab-analysis.

## Code Availability

R version 4.2.1 was used with the base packages and the following additional packages: stddiff, tidyverse, jtools, FSA, ggpubr, readxl, psych, and sjmisc. Custom scripts were used for the web application, data processing, and analysis and are open source at github.com/stanfordnmbl/motionlab-analysis.

## Author Contributions

M.B., L.K., J.H., and S.D conceptualized and designed the study. M.B. and L.K. conducted the study and performed data analysis. M.B. and L.K. wrote the first draft and all authors revised the manuscript. A.F. and S.U. conducted the motion capture experiments. All authors read and approved the final manuscript.

## Extended Data

**Extended Data Fig. 1.**
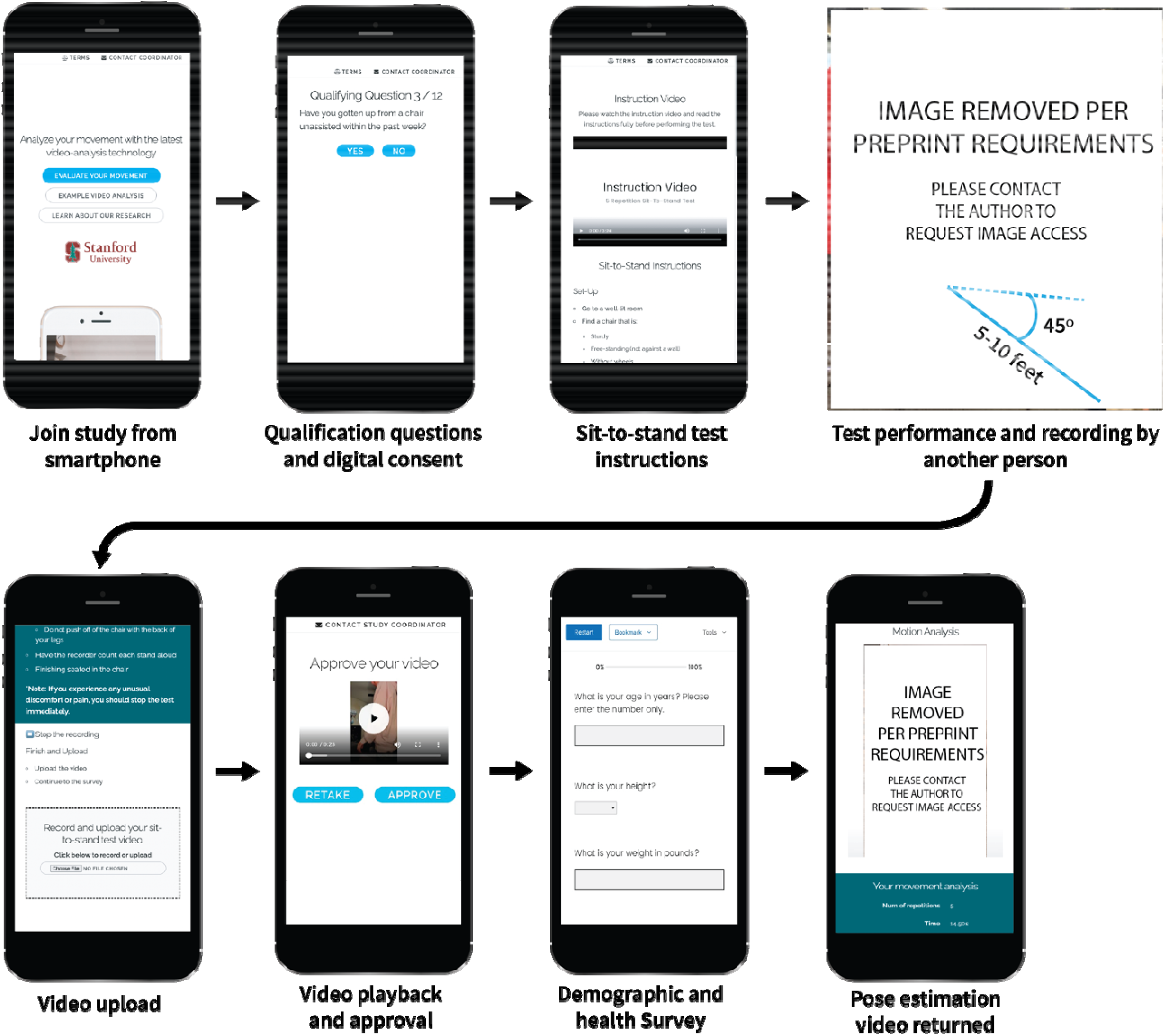
Step-by-step details of the user-facing side of the web application.

**Extended Data Fig. 2.**
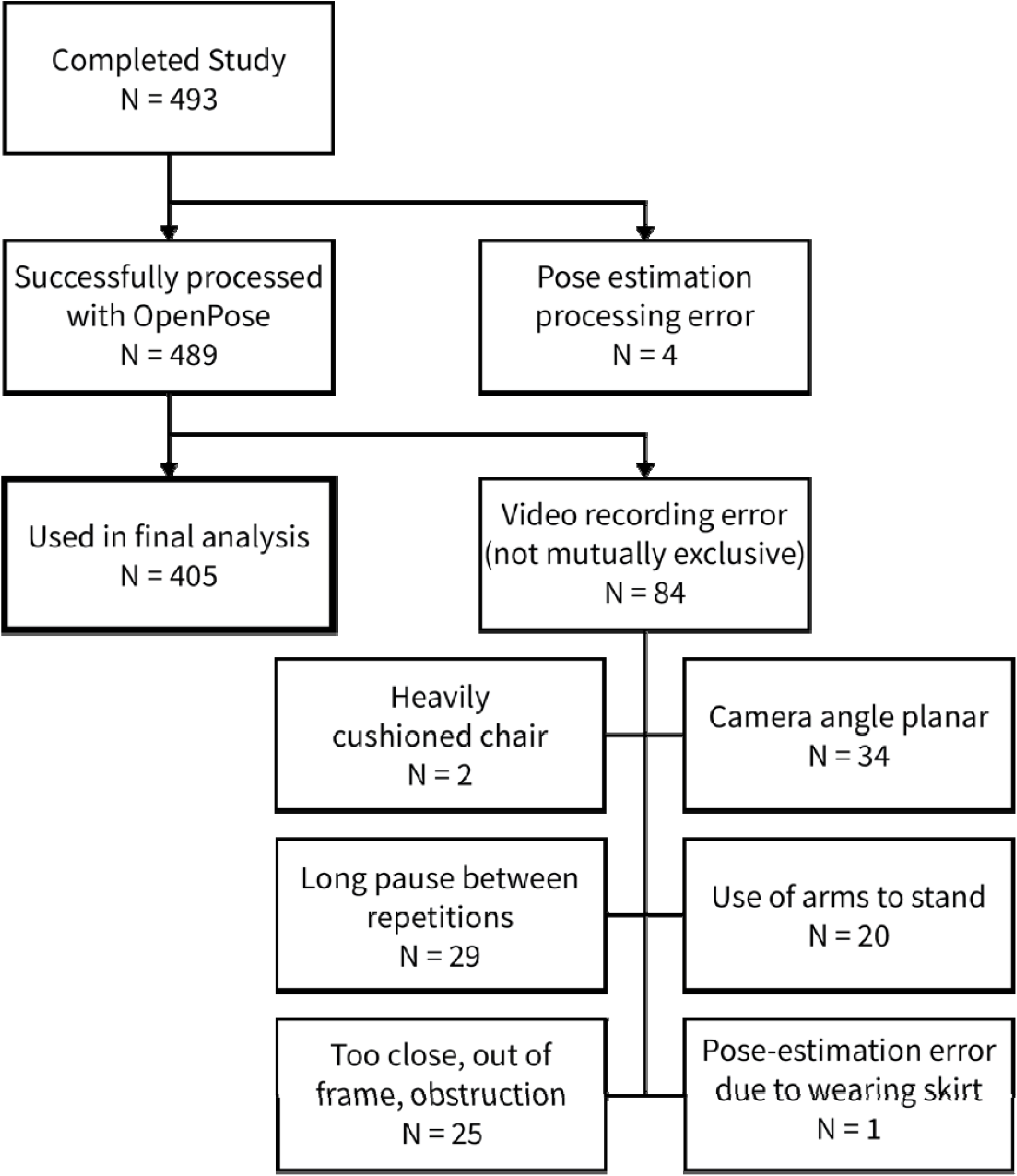
Flowchart of data included in the final analysis.

**Extended Data Fig. 3.**
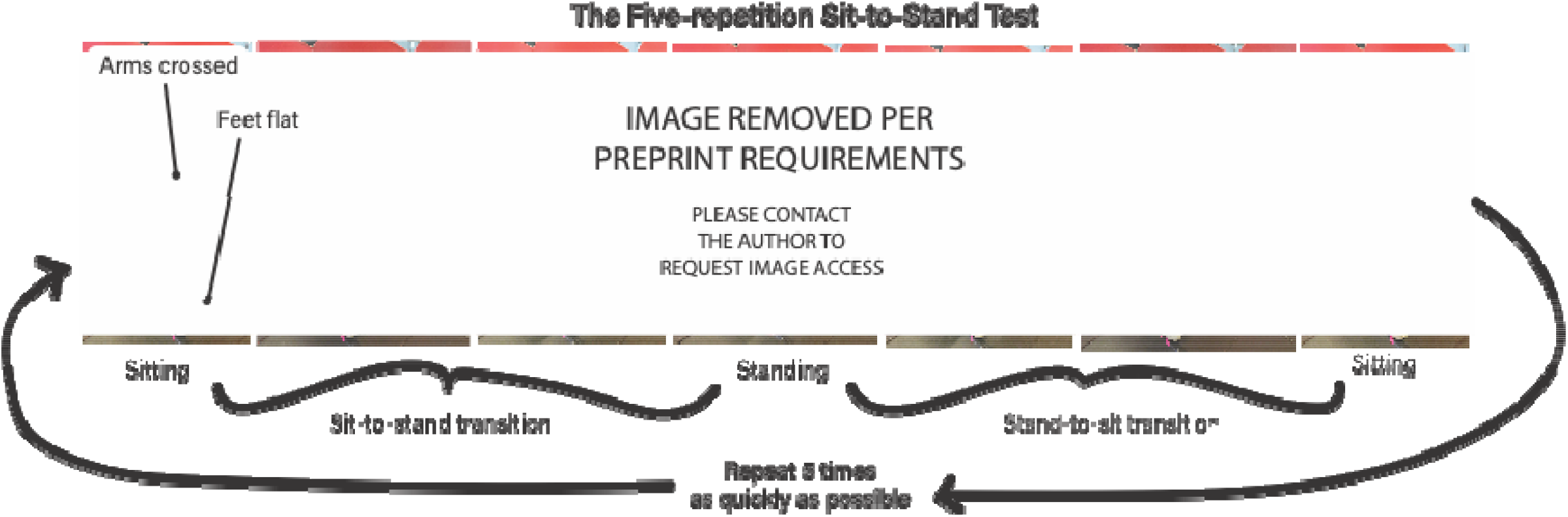
The Five-Repetition Sit-to-Stand Test. Individuals start sitting down with their arms crossed in front of their chest and their feet flat on the floor. They then rise to stand (sit-to-stand transition) and sit back down (stand-to-sit transition) five times as quickly as possible.

**Extended Data Table 1.** Participant characteristics.

| Variable | 18+ Years<br>N = 405 | 50+ Years<br>N = 106 |
| --- | --- | --- |
| **Participant Characteristics** |  |  |
| Age (years), mean ± SD (min, max) | 37 ± 18 (18, 96) | 64 ± 10 (50, 96) |
| Age binned, n (%)<sup>*</sup> |  |  |
| 18-29 years | 212 (52%) | - |
| 30-39 years | 62 (15%) | - |
| 40-49 years | 25 (6%) | - |
| 50-59 years | 44 (11%) | 44 (42%) |
| 60-69 years | 35 (9%) | 35 (33%) |
| 70 + years | 27 (7%) | 27 (25%) |
| Sex, n (%) |  |  |
| Male | 191 (47%) | 48 (55%) |
| Female | 214 (53%) | 58 (45%) |
| Gender, n (%) |  |  |
| Male | 192 (47%) | 48 (55%) |
| Female | 209 (52%) | 58 (45%) |
| Non-Binary | 2 (<1%) | 0 (0%) |
| Other | 1 (<1%) | 0 (0%) |
| Prefer not to answer | 1 (<1%) | 0 (0%) |
| Height (in), mean ± SD (min, max) | 67.3 ± 4.0 (55, 77) | 66.5 ± 3.8 (55, 75) |
| Body weight (lbs), mean ± SD (min, max) | 158 ± 34 (70, 280) | 164 ± 34 (70, 280) |
| Ethnicity, n (%) |  |  |
| White | 243 (60%) | 79 (75%) |
| Black or African American | 14 (3%) | 1 (1%) |
| Hispanic, Latino, or Spanish Origin | 16 (4%) | 2 (2%) |
| Asian | 103 (25%) | 19 (18%) |
| Mixed race | 24 (6%) | 2 (2%) |
| Other | 5 (1%) | 3 (3%) |
| Education, n (%) |  |  |
| Less than high school | 0 (0%) | 0 (0%) |
| High school graduate | 16 (4%) | 4 (4%) |
| Some college | 49 (12%) | 11 (10%) |
| College Degree or Higher | 338 (84%) | 89 (84%) |
| Prefer not to answer | 2 (<1%) | 2 (2%) |
| Employment, n (%) |  |  |
| Employed full time (40+ hours per week) | 190 (47%) | 46 (43%) |
| Employed part time (< 40 hours per week) | 25 (6%) | 9 (9%) |
| Student | 129 (32%) | 1 (1%) |
| Unemployed | 9 (2%) | 3 (3%) |
| Self-employed | 13 (3%) | 10 (9%) |
| Retired | 33 (8%) | 33 (31%) |
| Homemaker | 3 (<1%) | 2 (2%) |
| Prefer not to answer | 3 (<1%) | 2 (2%) |
| Income, n (%) |  |  |
| < \$10,000 | 17 (4%) | 0 (0%) |
| \$10,000 to \$24,999 | 12 (3%) | 0 (0%) |
| \$25,000 to \$49,999 | 53 (13%) | 4 (4%) |
| \$50,000 to \$99,999 | 98 (24%) | 25 (24%) |
| > \$100,000 | 153 (38%) | 48 (45%) |
| Prefer not to answer | 72 (18%) | 29 (27%) |
| State of residence (total #) | 35 | 25 |
| **Health Measures** |  |  |
| BMI (kg/m<sup>2</sup>), mean ± SD (min, max) | 24.3 ± 4.1 (16.3, 41.0) | 25.9 ± 4.5 (18.6, 40.2) |
| Physical health (T-score),<br>mean ± SD (min, max) | 54.9 ± 6.7 (32.4, 67.7) | 53.3 ± 7.1 (32.4, 67.7) |
| Mental health (T-score),<br>mean ± SD (min, max) | 53.0 ± 7.4 (28.4, 67.6) | 54.6 ± 7.0 (38.8, 67.6) |
| Osteoarthritis (Knee or Hip; Yes), n (%) | 30 (7%) | 29 (27%) |
| Hip | 8 (2%) | 8 (8%) |
| Knee | 26 (6%) | 25 (24%) |
| **5 STS Measures** |  |  |
| Total time (s) | 11.4 ± 3.4 (4.3, 32.9) | 13.2 ± 4.3 (5.0, 32.9) |
| Maximum trunk angle during sit-to-stand transition (deg) | 200.5 ± 8.3<br>(180.9, 239.3) | 201.9 ± 9.9<br>(180.9, 239.3) |
| Maximum trunk angular acceleration during sit-to-stand transition (deg/s<sup>2</sup>) | 685.9 ± 162.7<br>(297.3, 1279.4) | 678.7 ± 158.2<br>(297.3, 1216.4) |
| *Rounded percentages may result in sums slightly above or below 100%.<br>Abbreviations: BMI, Body mass index; Physical Health, PROMIS Global-10: Global Physical Health; Mental Health, PROMIS Global-10: Global Mental Health; |  |  |

**Extended Data Table 2.** Pearson correlations between measures and five-repetition sit-to-stand test time and kinematics for all participants (N = 405). P-values account for multiple comparisons using a false discovery rate for 21 comparisons.

|  | Time | Trunk Angle | Trunk<br>Angular<br>Acceleration |
| --- | --- | --- | --- |
| **Age** | 0.35**** | 0.15* | -0.02 |
| **Sex** | 0.03 | 0.00 | 0.05 |
| **BMI** | 0.20*** | 0.07 | -0.02 |
| **Ethnicity** | 0.05 | 0.16** | 0.01 |
| **Physical Health** | -0.20**** | -0.01 | -0.04 |
| **Mental Health** | 0.02 | 0.06 | -0.01 |
| **Osteoarthritis** | 0.18** | 0.18** | 0.06 |

\* $P < 0.05$
\*\* $P < 0.01$
\*\*\* $P < 0.001$
\*\*\*\* $P < 0.0001$
Abbreviations: BMI, Body mass index; Physical Health, PROMIS
Global-10: Global Physical Health; Mental Health, PROMIS Global-
10: Global Mental Health

**Extended Data Table 3.** Logistic regression analysis on a diagnosis of knee or hip osteoarthritis (N=405).

| **Dependent Variable** | **Independent Variables** | **$\beta$ (SE)** | **OR** | **(95% CI)** | **P** |
| --- | --- | --- | --- | --- | --- |
| Osteoarthritis<br>(yes/no = 1/0) | Age | 0.119 (0.012) | 1.126 | (0.094, 0.143) | <0.001 |
|  | Sex | -0.020 (0.023) | 0.980 | (-0.066, 0.026) | 0.393 |
|  | BMI | 0.008 (0.012) | 1.008 | (-0.016, 0.031) | 0.528 |
|  | Time | 0.001 (0.012) | 0.999 | (-0.025, 0.024) | 0.957 |
|  | Trunk Angle | 0.029 (0.012) | 1.030 | (0.007, 0.052) | 0.012 |

Abbreviations: BMI, Body mass index

**Extended Data Table 4.**
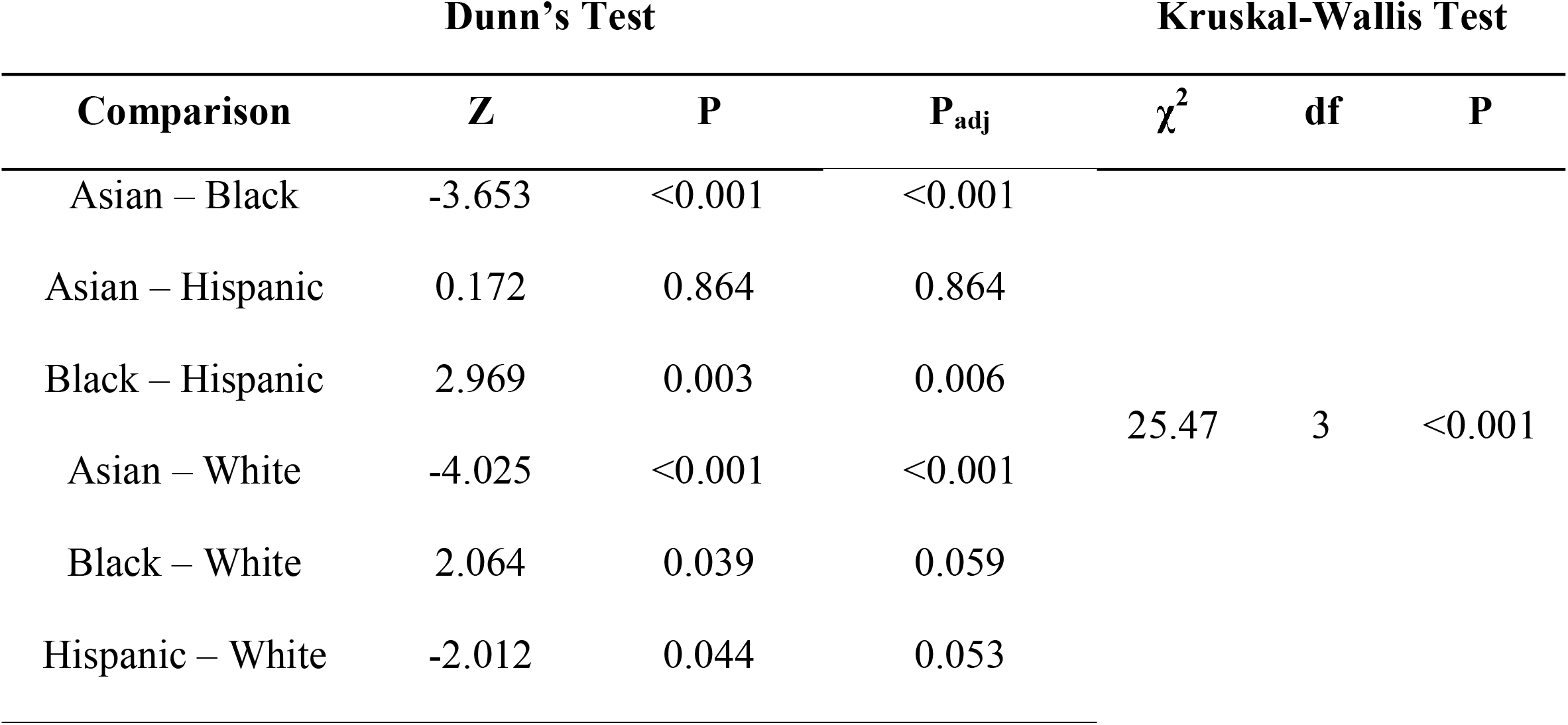
Results of a Kruskal-Wallis rank sum test and posthoc Dunn’s test for differences in trunk angle between races and ethnicities (N=405). P-values account for multiple comparisons using a false discovery rate for 6 comparisons.

**Extended Data Table 5.** Logistic regression analysis on race (N_White_=243; N_Asian_=103).

| **Dependent Variable** | **Independent Variables** | **B (SE)** | **OR** | **(95% CI)** | **P** |
| --- | --- | --- | --- | --- | --- |
| Ethnicity<br>(White = 0;<br>Asian = 1) | Age | -0.043 (0.026) | 0.958 | (-0.094, 0.007) | 0.093 |
|  | Sex | 0.013 (0.047) | 1.013 | (-0.079, 0.105) | 0.781 |
|  | BMI | -0.114 (0.024) | 0.892 | (-0.162, -0.066) | <0.001 |
|  | Time | -0.018 (0.025) | 0.982 | (-0.068, 0.031) | 0.467 |
|  | Physical Health | -0.086 (0.024) | 0.918 | (-0.132, -0.039) | <0.001 |
|  | Trunk Angle | -0.084 (0.023) | 0.920 | (-0.130, -0.038) | <0.001 |

Abbreviations: BMI, Body mass index; Physical Health, PROMIS Global-10: Global Physical Health

**Extended Data Table 6.** Pearson correlations between measures and five-repetition sit-to-stand test time and kinematics for participants 50 years of age or older (N = 106). P-values account for multiple comparisons by controlling for the false discovery rate for 21 comparisons.

|  | Time | Trunk Angle | Trunk<br>Angular<br>Acceleration |
| --- | --- | --- | --- |
| **Physical Health** | -0.39*** | -0.21 | 0.09 |
| **Mental Health** | -0.06 | 0.09 | 0.28* |

\* $P < 0.05$
\*\* $P < 0.01$
\*\*\* $P < 0.001$
\*\*\*\* $P < 0.0001$
Abbreviations: BMI, Body mass index; Physical Health, PROMIS
Global-10: Global Physical Health; Mental Health, PROMIS Global-
10: Global Mental Health

**Extended Data Table 7.** Linear regression analysis on mental health score for individuals 50 years of age or older (N=106).

| **Dependent Variable** | **Independent Variables** | **$\beta$** | **(95% CI)** | **P** | **Adj. R<sup>2</sup>** | **F (P)** |
| --- | --- | --- | --- | --- | --- | --- |
| Mental Health (T-Score) | Age | 0.803 | (-0.632, 2.238) | 0.270 | 0.108 | 3.544 (0.005) |
|  | Sex | -1.874 | (-4.492, 0.744) | 0.159 |  |  |
|  | BMI | -1.539 | (-2.898, -0.180) | 0.027 |  |  |
|  | Time | -0.136 | (-1.569, 1.297) | 0.851 |  |  |
|  | Trunk Angular Acceleration | 1.705 | (0.376, 3.034) | 0.012 |  |  |

Abbreviations: BMI, Body mass index

## References

1. The World Health Organization Quality of Life assessment (WHOQOL): position paper from the World Health Organization. Soc. Sci. Med. 41, 1403–1409 (1995).

2. Sibbritt, D. W., Byles, J. E. & Regan, C. Factors associated with decline in physical functional health in a cohort of older women. Age Ageing 36, 382–388 (2007).

3. Araujo, I. L. A., Castro, M. C., Daltro, C. & Matos, M. A. Quality of Life and Functional Independence in Patients with Osteoarthritis of the Knee. Knee Surgery & Related Research vol. 28 219–224 (2016).

4. van Lummel, R. C. et al. The Instrumented Sit-to-Stand Test (iSTS) Has Greater Clinical Relevance than the Manually Recorded Sit-to-Stand Test in Older Adults. PLoS One 11, e0157968 (2016).

5. Shukla, B., Bassement, J., Vijay, V., Yadav, S. & Hewson, D. Instrumented Analysis of the Sit-to-Stand Movement for Geriatric Screening: A Systematic Review. Bioengineering (Basel) 7, (2020).

6. Turcot, K., Armand, S., Fritschy, D., Hoffmeyer, P. & Suvà, D. Sit-to-stand alterations in advanced knee osteoarthritis. Gait Posture 36, 68–72 (2012).

7. Millor, N., Lecumberri, P., Gomez, M., Martinez-Ramirez, A. & Izquierdo, M. Kinematic parameters to evaluate functional performance of sit-to-stand and stand-to-sit transitions using motion sensor devices: a systematic review. IEEE Trans. Neural Syst. Rehabil. Eng. 22, 926–936 (2014).

8. Pourahmadi, M. R. et al. Kinematics of the Spine During Sit-to-Stand Movement Using Motion Analysis Systems: A Systematic Review of Literature. Journal of Sport Rehabilitation vol. 28 77–93 (2019).

9. Silver, L. & Taylor, K. Smartphone ownership is growing rapidly around the world, but not always equally. Pew Research Center. (2019).

10. Toshev, A. & Szegedy, C. DeepPose: Human Pose Estimation via Deep Neural Networks. 2014 IEEE Conference on Computer Vision and Pattern Recognition (2014).

11. Pishchulin, L. et al. DeepCut: Joint Subset Partition and Labeling for Multi Person Pose Estimation. 2016 IEEE Conference on Computer Vision and Pattern Recognition (CVPR) (2016).

12. Cao, Z., Hidalgo, G., Simon, T., Wei, S.-E. & Sheikh, Y. OpenPose: Realtime Multi-Person 2D Pose Estimation using Part Affinity Fields. arXiv [cs.CV] (2018).

13. Mathis, A. et al. DeepLabCut: markerless pose estimation of user-defined body parts with deep learning. Nat. Neurosci. 21, 1281–1289 (2018).

14. Kidziński, Ł. et al. Deep neural networks enable quantitative movement analysis using single-camera videos. Nat. Commun. 11, 4054 (2020).

15. Stenum, J., Rossi, C. & Roemmich, R. T. Two-dimensional video-based analysis of human gait using pose estimation. PLoS Comput. Biol. 17, e1008935 (2021).

16. Ejupi, A. et al. Kinect-Based Five-Times-Sit-to-Stand Test for Clinical and In-Home Assessment of Fall Risk in Older People. Gerontology 62, 118–124 (2015).

17. O’Neill, T. W., McCabe, P. S. & McBeth, J. Update on the epidemiology, risk factors and disease outcomes of osteoarthritis. Best Pract. Res. Clin. Rheumatol. 32, 312–326 (2018).

18. Loureiro, A., Mills, P. M. & Barrett, R. S. Muscle weakness in hip osteoarthritis: a systematic review. Arthritis Care Res. 65, 340–352 (2013).

19. Higgs, J. P. et al. Individuals with mild-to-moderate hip osteoarthritis exhibit altered pelvis and hip kinematics during sit-to-stand. Gait Posture 71, 267–272 (2019).

20. Mullineaux, D. R., Bartlett, R. M. & Bennett, S. Research design and statistics in biomechanics and motor control. J. Sports Sci. 19, 739–760 (2001).

21. Doorenbosch, C. A. M., Harlaar, J., Roebroeck, M. E. & Lankhorst, G. J. Two strategies of transferring from sit-to-stand; The activation of monoarticular and biarticular muscles. J. Biomech. 27, 1299–1307 (1994).

22. Uhlrich, S. D. et al. OpenCap: 3D human movement dynamics from smartphone videos. bioRxiv 2022.07.07.499061 (2022).

23. Lord, S. R., Murray, S. M., Chapman, K., Munro, B. & Tiedemann, A. Sit-to-stand performance depends on sensation, speed, balance, and psychological status in addition to strength in older people. J. Gerontol. A Biol. Sci. Med. Sci. 57, M539–43 (2002).

24. Bohannon, R. W., Bubela, D. J., Magasi, S. R., Wang, Y.-C. & Gershon, R. C. Sit-to-stand test: Performance and determinants across the age-span. Isokinet. Exerc. Sci. 18, 235–240 (2010).

25. Janssen, W. G. M., Bussmann, H. B. J. & Stam, H. J. Determinants of the sit-to-stand movement: A review. Phys. Ther. 82, 866–879 (2002).

26. Jordan, J. M. et al. Ethnic health disparities in arthritis and musculoskeletal diseases: report of a scientific conference. Arthritis Rheum. 46, 2280–2286 (2002).

27. Hill, C. N., Reed, W., Schmitt, D., Sands, L. P. & Queen, R. M. Racial differences in gait mechanics. J. Biomech. 112, 110070 (2020).

28. Lim, D., Norman, R. & Robinson, S. Consumer preference to utilise a mobile health app: A stated preference experiment. PLoS One 15, 1–12 (2020).

29. Andriluka, M., Pishchulin, L., Gehler, P. & Schiele, B. 2D human pose estimation: New benchmark and state of the art analysis. Proceedings of the IEEE Computer Society Conference on Computer Vision and Pattern Recognition 3686–3693 (2014).

30. Lin, T. Y. et al. Microsoft COCO: Common objects in context. in European Conference on Cumputer Vision 740–755 (2014).

31. Warburton, Jamnik & Bredin. The 2020 Physical Activity Readiness Questionnaire for Everyone (PAR-Q+) and electronic Physical Activity Readiness Medical Examination (ePARmed-X+): 2020 …. The Health & Fitness.

32. Austin, P. C. An Introduction to Propensity Score Methods for Reducing the Effects of Confounding in Observational Studies. Multivariate Behav. Res. 46, 399–424 (2011).

33. Stuart, E. A., Lee, B. K. & Leacy, F. P. Prognostic score-based balance measures can be a useful diagnostic for propensity score methods in comparative effectiveness research. J. Clin. Epidemiol. 66, S84–S90.e1 (2013).

34. Rw, B. Alternatives for measuring knee extension strength of the elderly at home. Clin. Rehabil. 12, 434–440 (1998).

35. Riley, P. O., Schenkman, M. L., Mann, R. W. & Hodge, W. A. Mechanics of a constrained chair-rise. J. Biomech. 24, 77–85 (1991).

36. Rees-Punia, E., Rittase, M. H. & Patel, A. V. A method for remotely measuring physical function in large epidemiologic cohorts: Feasibility and validity of a video-guided sit-to-stand test. PLoS One 16, e0260332 (2021).

37. Bohannon, R. W. Reference values for the five-repetition sit-to-stand test: a descriptive meta-analysis of data from elders. Percept. Mot. Skills 103, 215–222 (2006).

38. Newcomer, K. L., Krug, H. E. & Mahowald, M. L. Validity and reliability of the timed-stands test for patients with rheumatoid arthritis and other chronic diseases. J. Rheumatol. 20, 21–27 (1993).

39. Jones, S. E. et al. The five-repetition sit-to-stand test as a functional outcome measure in COPD. Thorax 68, 1015–1020 (2013).

40. Duncan, R. P., Leddy, A. L. & Earhart, G. M. Five times sit-to-stand test performance in Parkinson’s disease. Arch. Phys. Med. Rehabil. 92, 1431–1436 (2011).

41. Staartjes, V. E. & Schröder, M. L. The five-repetition sit-to-stand test: evaluation of a simple and objective tool for the assessment of degenerative pathologies of the lumbar spine. J. Neurosurg. Spine 29, 380–387 (2018).

42. Barile, J. P. et al. Monitoring population health for Healthy People 2020: evaluation of the NIH PROMIS® Global Health, CDC Healthy Days, and satisfaction with life instruments. Qual. Life Res. 22, 1201–1211 (2013).

43. Hays, R. D., Bjorner, J. B., Revicki, D. A., Spritzer, K. L. & Cella, D. Development of physical and mental health summary scores from the patient-reported outcomes measurement information system (PROMIS) global items. Qual. Life Res. 18, 873–880 (2009).

44. pose-estimation. GitHub Topics https://github.com/topics/pose-estimation.

45. Mroz, S. et al. Comparing the Quality of Human Pose Estimation with BlazePose or OpenPose. in 2021 4th International Conference on Bio-Engineering for Smart Technologies (BioSMART) 1–4 (2021).

46. Zhang, F. et al. Comparison of OpenPose and HyperPose artificial intelligence models for analysis of hand-held smartphone videos. in 2021 IEEE International Symposium on Medical Measurements and Applications (MeMeA) 1–6 (2021).

47. Benjamini, Y. & Hochberg, Y. Controlling the False Discovery Rate: A Practical and Powerful Approach to Multiple Testing. J. R. Stat. Soc. Series B Stat. Methodol. 57, 289–300 (1995).

48. Delp, S. L. et al. OpenSim: open-source software to create and analyze dynamic simulations of movement. IEEE Trans. Biomed. Eng. 54, 1940–1950 (2007).

49. Seth, A. et al. OpenSim: Simulating musculoskeletal dynamics and neuromuscular control to study human and animal movement. PLoS Comput. Biol. 14, e1006223 (2018).

50. Lai, A. K. M., Arnold, A. S. & Wakeling, J. M. Why are Antagonist Muscles Co-activated in My Simulation? A Musculoskeletal Model for Analysing Human Locomotor Tasks. Ann. Biomed. Eng. 45, 2762–2774 (2017).

